# Advancing *Legionella pneumophila* genomic surveillance with a high-resolution cg/wgMLST schema for outbreak detection and investigation

**DOI:** 10.64898/2026.02.18.26346554

**Authors:** Verónica Mixão, Christophe Ginevra, Camille Jacqueline, Sophie Jarraud, Marco Gabrielli, João Paulo Gomes, Melisa J. Willby, Jennafer A. P. Hamlin, Vítor Borges

## Abstract

**Introduction:** Sequence-based typing (SBT) has been the standard molecular typing method for understanding *Legionella pneumophila* genetic relationships. However, genome-scale typing approaches, namely core-genome (cg) or whole-genome (wg) multilocus sequence typing (MLST), provide higher discriminatory power. To advance these capabilities, the *Legionella* International Typing (LIT) workgroup was established to develop, evaluate, and disseminate a novel cgMLST schema with enhanced wgMLST resolution for *L. pneumophila* investigations.

**Methods:** We created and populated the LIT cg/wgMLST schema with chewBBACA software using more than 9000 genome assemblies representative of the species diversity. We applied a multi-step refinement workflow, considering loci prevalence, diversity and presence/absence profile across the species tree, to select the final cg/wgMLST loci, and compared the performance of the LIT cgMLST schema with the previously used 1521-loci schema and assessed its congruence with SBT.

**Results:** The LIT schema includes 2009 loci present in 98% of the dataset, forming the static cgMLST schema for routine genomic surveillance, plus 2698 accessory loci for an in-depth wgMLST analysis of clusters of interest. The LIT cgMLST schema maintains moderate agreement with SBT and presents high clustering congruence with the 1521-loci schema, while providing increased resolution. Analysis of epidemiologically related isolates using the LIT cgMLST schema for initial cluster delineation, followed by cluster-specific dynamic wgMLST analysis extending the cgMLST with accessory loci shared among isolates within each cluster, demonstrated increased confidence for outbreak investigation and source identification.

**Conclusions:** The LIT schema is expected to contribute to harmonizing genomic surveillance of Legionnaires’ disease at both local and global levels. The schema and associated resources for local implementation are available on Zenodo (https://doi.org/10.5281/zenodo.17871973).

## Introduction

The gram-negative bacterium *Legionella pneumophila* causes Legionnaires’ disease (LD), a severe pneumonia [1]. Although *L. pneumophila* is found in natural habitats (e.g. lakes and soil), it can also proliferate in human-made systems such as cooling towers, plumbing systems, and hot tubs, particularly when poorly maintained [1]. Infection occurs when aerosols from water sources containing *L. pneumophila* are inhaled [1]. As a result, LD presents predominantly as isolated, sporadic cases but can also occur as outbreaks when individuals share exposure to a common source within a defined timeframe [1]. Surveillance data from the U.S. Centers for Disease Control and Prevention (CDC) and the European Centre for Disease Prevention and Control (ECDC) show an increasing incidence of LD [2,3], and a recent summary of several large outbreaks that occurred in 2024, emphasizes the need for heightened clinical and public health awareness [4]. Furthermore, outbreaks are costly. For example, in New Zealand, LD cost an estimated NZ$2.1 million annually during 2016-2020, based on an average of 143 hospitalized cases with a median stay of six days [5].

Multi-locus sequence typing (MLST) is the most widely used method for bacterial characterization [6]. In *L. pneumophila*, MLST is known as sequence-based typing (SBT), which targets seven genes to define sequence type (ST). Developed in the early 2000s by the European Society of Clinical Microbiology and Infectious Diseases (ESCMID) Study Group for *Legionella* Infections (ESGLI), SBT by Sanger sequencing has been the standard method for over two decades for understanding genetic relatedness [7]. However, this technique is labor-intensive, low throughput, and may lack sufficient resolution [8–11], thus, highlighting the need for Whole-Genome Sequencing (WGS)-based methods that capture broader genomic variation. Indeed, *L. pneumophila* presents substantial genetic diversity within and between STs (e.g. ST1), and recombination shapes its genomic landscape [8–14]. Although single-nucleotide polymorphism (SNP)-based analyses provide high discriminatory power between closely related isolates and have been crucial in numerous *L. pneumophila* outbreak investigations [15–17], their application in large and diverse datasets/collections remains challenging and often entails a decrease in resolution [14,18]. Core-genome MLST (cgMLST) approaches, which offer a high discriminatory power, while inherently buffering the noise introduced by recombination, are a good alternative for *L. pneumophila* routine genomic surveillance. In 2015, Moran-Gilad *et al*. (2015) published the *L. pneumophila* 1521-loci cgMLST schema based on limited diversity (38 genome sequences) [8]. Later on, David *et al*. (2016) proposed a 50-gene alternative relying on a larger dataset (335 genomes), in an attempt to balance resolution with epidemiological concordance [9]. Although these schemas have clarified outbreaks where SBT lacked sufficient discrimination [14,19], they have not replaced SBT, mainly due to dependence on multiple commercial platforms and focus on local deployment hampering the standardized application at the global level [9,20]. Furthermore, these cgMLST schemas lack sufficient discriminatory power for investigating certain *L. pneumophila* clusters, where isolates’ genomes are frequently nearly identical at the core-genome level [21]. We reasoned that a publically available typing schema enabling a dynamic wgMLST approach could overcome these limitations. This strategy combines an initial large-scale high-resolution cgMLST analysis with a subsequent in-depth cluster zoom-in [22,23] and has already shown promising results for *Salmonella enterica*, *Escherichia coli* and *Campylobacter jejuni* [24], making it potentially suited to *L. pneumophila* genomic characterization.

In this context, the *Legionella* International Typing (LIT) workgroup was established to develop, evaluate, and disseminate a novel cgMLST schema with enhanced wgMLST resolution for *L. pneumophila*. This schema is expected to improve routine surveillance and outbreak discrimination, offering a robust foundation for consistent genomic characterization and future descriptive nomenclature development in *L. pneumophila*.

## Methods

### Dataset selection and curation

To create a novel *L. pneumophila* cg/wgMLST schema, a dataset of genome assemblies’ representative of the species diversity was compiled. All *L. pneumophila* genome assemblies available in the NCBI database on May, 30^th^ 2024 (n=5722) were downloaded. These assemblies were supplemented with the genome assemblies of all *L. pneumophila* isolates (n=4499) from the National Reference Centre of *Legionella* (NRCL) of the Institute of Infectious Agents (Lyon, France) that were collected up to the same date. This dataset was curated by removing contigs with <500bp and excluding assemblies with abnormal genome size (diverging from 3.5Mbp ±20%) or which were duplicated (*i.e.*, same isolate sequenced twice in the NRCL dataset). Furthermore, fastANI v1.33 [25] was used for species classification using an in-house database containing one representative genome per subspecies, and the assemblies that were not assigned as *L. pneumophila* were also excluded (<95% identity). The final curated dataset comprised 9657 genome assemblies (details in Supplementary file S1).

### Selection of high-quality assemblies for schema creation

After compiling the curated dataset, its genetic diversity was evaluated using a previously developed cgMLST schema, hereinafter referred to as the 1521-loci schema [8]. The 1521-loci schema was adapted for local deployment with chewBBACA software (https://github.com/B-UMMI/chewBBACA) [26] using chewBBACA v3.6 PrepExternalSchema module with the *L. pneumophila* training file available with the tool. Allele calling for the 9657 assemblies was then performed with chewBBACA v3.6 AlleleCall module and was repeated until no new alleles were inferred (3 rounds). The resulting allele matrix was provided as input to ReporTree v2.6 [23] to identify genetic clusters for all possible allelic distance thresholds, excluding isolates with less than 90% loci called, and using the single-linkage hierarchical clustering (HC) algorithm. Clustering information, together with assembly metrics, was provided as input to DownTree v1.0.0 (https://github.com/insapathogenomics/ACDTree/tree/main/DownTree) [27]. High-quality assemblies (meeting a criteria of ≤120 contigs) were selected, when applicable, from every singleton/cluster defined at 4 allelic differences (ADs), which is the threshold commonly used to identify potential *L. pneumophila* outbreak clusters with the 1521-loci schema [8,20], up to a maximum of three assemblies per cluster. For larger clusters, the three most genetically divergent assemblies were selected, according to the DownTree method. This resulted in 4403 high-quality assemblies selected to capture species’ overall diversity for schema creation, representing 95% of the 3086 genetic groups detected at 4 ADs and ensuring 98% sample representativeness (Supplementary figure S1 and Supplementary file S1).

### Schema creation, loci quality filtering and nomenclature

The novel schema was created with chewBBACA v3.6 [26] CreateSchema module, providing the selected 4403 representative assemblies as input. This resulted in an initial *L. pneumophila* cg/wgMLST schema with 10482 loci. Allele calling for the 9657 assemblies was then performed with chewBBACA v3.6 [26] AlleleCall module and repeated until no new alleles were inferred (12 rounds). Next, PanTree v1.0.0 (https://github.com/insapathogenomics/ACDTree/tree/main/PanTree) [27] was used to obtain information about locus diversity, prevalence in the dataset and presence/absence across the different genetic clusters. PanTree takes as input the allelic matrix of the novel schema, along with the ReporTree-derived clustering information for the 9657 assemblies based on the 1521-loci schema, including all AD threshold-level partitions and the corresponding dendrogram [8]. Based on PanTree results, non-discriminative (*i.e.,* with only one allele) and low prevalent (*i.e.*, called in less than 1% of the 4 AD clusters) loci were excluded. Moreover, loci identified by chewBBACA as paralogs, and those with an allele size mode outside the ±20% “size-threshold” (flagged as ALM/ASM) and/or with a non-informative paralogous hit (flagged as NIPH/NIPHM) in at least 10% of the isolates were also removed. Additionally, loci corresponding to putative contamination, identified with Kraken2 [28] using the PlusPF database (downloaded in April 2021), were excluded. This resulted in a final wgMLST schema with 4707 loci (Supplementary file S2).

For locus nomenclature, BLASTn [29] was performed between the alleles obtained for the *L. pneumophila* Philadelphia strain and the open reading frames (ORFs) of its reference genome (accession number: NC_002942.5). Loci with an ORF match (100% identity) were sequentially named according to their position in the reference genome, from lpLIT0001 to lpLIT2552 (where lp stands for *Legionella pneumophila* and LIT for *Legionella* International Typing). The remaining loci were named according to their decreasing prevalence in the 9657 assemblies dataset, from lpLIT2553 to lpLIT4707 (Supplementary file S2). Finally, chewBBACA v3.6 PrepExternalSchema module was run to prepare the schema for the “schema population” step.

### cgMLST loci selection and “schema population”

For the selection of the static cgMLST loci, allele calling was performed for the 9657 assemblies using the chewBBACA v3.6 [26] AlleleCall module with the 4707-loci wgMLST schema until no new alleles were inferred (20 rounds). For the selection of the static cgMLST schema, the resulting matrix was provided to ReporTree v2.6 [23] to identify genetic clusters at all possible allele distance thresholds using HC algorithm. Genetic clustering was then evaluated at four different loci presence thresholds: 95% (cgMLST 95), 98% (cgMLST 98), 99% (cgMLST 99) and 100% (cgMLST 100, strict core) of the dataset, using the *--site-inclusion* argument of ReporTree (Supplementary file S3). In each analysis, samples with less than 95% loci called were removed. Clustering congruence and resolution differences among the four generated cgMLST schemas were evaluated with EvalTree v1.0.0 (https://github.com/insapathogenomics/ACDTree/tree/main/EvalTree) [27]. Clustering congruence measures the similarity of isolate clustering across different cgMLST/wgMLST schemas, using the previously developed congruence score (CS) [24], which ranges from 0 (no congruence) to 3 (absolute congruence). Resolution reflects each schema’s ability to distinguish genetically distinct isolates at the highest resolution, here assessed as the number of identified genetic groups at 0 ADs. Based on these analyses, the final static cgMLST loci list was defined based on the 98% loci presence threshold (Supplementary file S2). The original 4707-loci schema (*i.e.,* prepared after the application of loci nomenclature) was then populated with the assemblies that had at least 95% loci called in this cgMLST (Supplementary file S1), through several rounds of chewBBACA v3.6 AlleleCall module until no new alleles were inferred (8 rounds). This procedure produced the final version of the novel cg/wgMLST schema, hereafter designated as the LIT schema, which is composed of 4707 loci, of which 2009 constitute the static cgMLST schema.

### *In silico* traditional typing

To ensure that SBT information was available for comparison with the LIT schema, *in silico* SBT was performed with el_gato v1.22.0 (https://github.com/CDCgov/el_gato) for the NCBI genome dataset (n=5708). For this analysis, the corresponding reads were used as input instead of assembled genomes, when available. The NRCL dataset was typed using the mompS tool (https://github.com/bioinfo-core-BGU/mompS) [30] with default parameters. SBT information is provided in Supplementary file S1.

### Congruence with the 1521-loci cgMLST schema and compatibility with SBT

To assess cluster congruence between the different typing pipelines, the same strategy used to define the static cgMLST (see “cgMLST loci selection and schema population” section), originally developed in a large-scale comparison of bacterial surveillance pipelines [24], was applied. Using the ReporTree v2.6 [23] outputs for each cgMLST schema as input, EvalTree v1.0.0 [27] evaluates clustering behavior across all possible thresholds and identifies cluster stability regions (*i.e.*, threshold ranges that generate consistent clustering patterns). EvalTree v1.0.0 also performs inter-pipeline clustering congruence analysis at all possible allelic distance threshold levels, identifying the thresholds that yield the most similar clustering results between pipelines (*i.e*., inter-pipeline corresponding points), defined as the threshold comparisons producing the highest CS values, considering only CS ≥2.85.

Cluster congruence was evaluated between the single-linkage hierarchical clustering of the 1521-loci schema and the LIT cgMLST schema, using a dataset composed of isolates passing the “loci called” criteria in both pipelines (90% and 95%, respectively) (Supplementary file S4). To assess the applicability of the LIT schema with different clustering algorithms, clustering congruence between HC and GrapeTree MSTreeV2 [31] was also evaluated. Additionally, EvalTree was used to determine the threshold level of the LIT cgMLST schema showing the best clustering congruence with SBT classifications, considering only samples for which an ST could be assigned (see section “*In sílico* traditional typing”). Clustering information obtained at all possible threshold levels with ReporTree are compiled in Supplementary files S3, S4 and S5, while EvalTree results are compiled in Supplementary file S6. For the ten most represented STs in the dataset (Supplementary file S1), the lowest threshold level at which all samples of the same ST clustered together in the LIT cgMLST schema (Supplementary file S7) was identified using the script *poli_typing.py* (available at https://github.com/insapathogenomics/WGS_cluster_congruence), as previously described [24].

### Application of the dynamic wgMLST approach to support outbreak investigation and source attribution

To assess the value of a dynamic wgMLST approach in *L. pneumophila* outbreak investigation, ReporTree v2.6 [23] was run on the entire wgMLST allelic matrix (9657 samples and 4707 loci) using the “zoom-clusters” mode. In this mode, after an initial clustering based on the static LIT cgMLST schema (loci list provided as input, https://github.com/insapathogenomics/ReporTree/blob/main/useful_loci_lists/cgMLST_Legionella_pneumophila_LIT.txt), ReporTree automatically performs a cluster-specific dynamic wgMLST analysis by extending the static schema with accessory loci shared by all isolates of the same cluster (*--site-inclusion* 1.0). The analysis was performed at the 6 ADs threshold level, which was identified as the value with the highest clustering congruence with the commonly used 4 ADs threshold used in the 1521-loci schema (see Results, Supplementary file S6). The results of this large-scale application of a dynamic wgMLST approach are compiled in Supplementary file S8.

To further illustrate the added value of the dynamic wgMLST approach in real-life *L. pneumophila* surveillance scenarios, nine groups of epidemiologically-related isolates (hereinafter designated as “investigations”) from the NRCL dataset (Supplementary file S8) were analyzed to assess improvements in resolution for both the identification of an outbreak and discrimination among isolates within each investigation. To this end, ReporTree v2.6 [23] was re-run with “zoom-cluster-of-interest” mode, requesting a dynamic wgMLST analysis only for clusters including at least one clinical sample (*--sample_of_interest*) for each of the nine investigations. The gains provided by the dynamic wgMLST approach relative to the initial static cgMLST clustering were evaluated based on four metrics: i) cluster delineation, defined as the ability to detect a (sub)cluster exclusively formed by all same-investigation isolates; ii) cluster separation, defined as the allelic distance of the shortest connection between investigation isolates and the other isolates of the cluster (if applicable); iii) cluster discrimination, defined as the number of non-redundant allelic profiles observed among isolates within each investigation; and iv) cluster confidence, defined as the strength of genetic evidence based on gains in at least one of the previous three metrics and/or in the number of loci supporting the investigation results.

For comparison, a reference-based SNP calling analysis was performed for each of the nine groups of epidemiologically-related isolates. Briefly, within each investigation, Trimmomatic v0.39 [32] was applied to trim reads of all the investigation-associated isolates. Then, Snippy v4.6.0 (https://github.com/tseemann/snippy), with default settings, was used for mapping and variant calling of all trimmed reads, with one clinical isolate selected as reference for each investigation.

## Results

### Novel LIT cg/wgMLST schema for monitoring *Legionella pneumophila* diversity and outbreak investigation

To develop a novel *L. pneumophila* cg/wgMLST schema (hereafter referred to as the LIT schema), a curated dataset with 9657 *L. pneumophila* genome assemblies was compiled, and a comprehensive and systematic multi-step methodology was applied (Figure 1). First, 4403 high-quality assemblies, representative of the species diversity, were selected for schema creation using chewBBACA [26] (Supplementary file S1). The initial draft schema, comprising 10482 loci, was subjected to a comprehensive curation process, resulting in a final LIT wgMLST schema with 4707 loci (Figure 1), with different prevalences in the global dataset (Figure 2A and Supplementary file S2). Around half of the loci are present in >80% of the genomes (Figure 2B and Supplementary file S2), with the remaining ones frequently displaying presence profiles that are specific to certain branch(es) of the species tree (Figure 2A). Each genome has a median of 2648 loci called (min=2343; max=2879).

**Figure 1.**
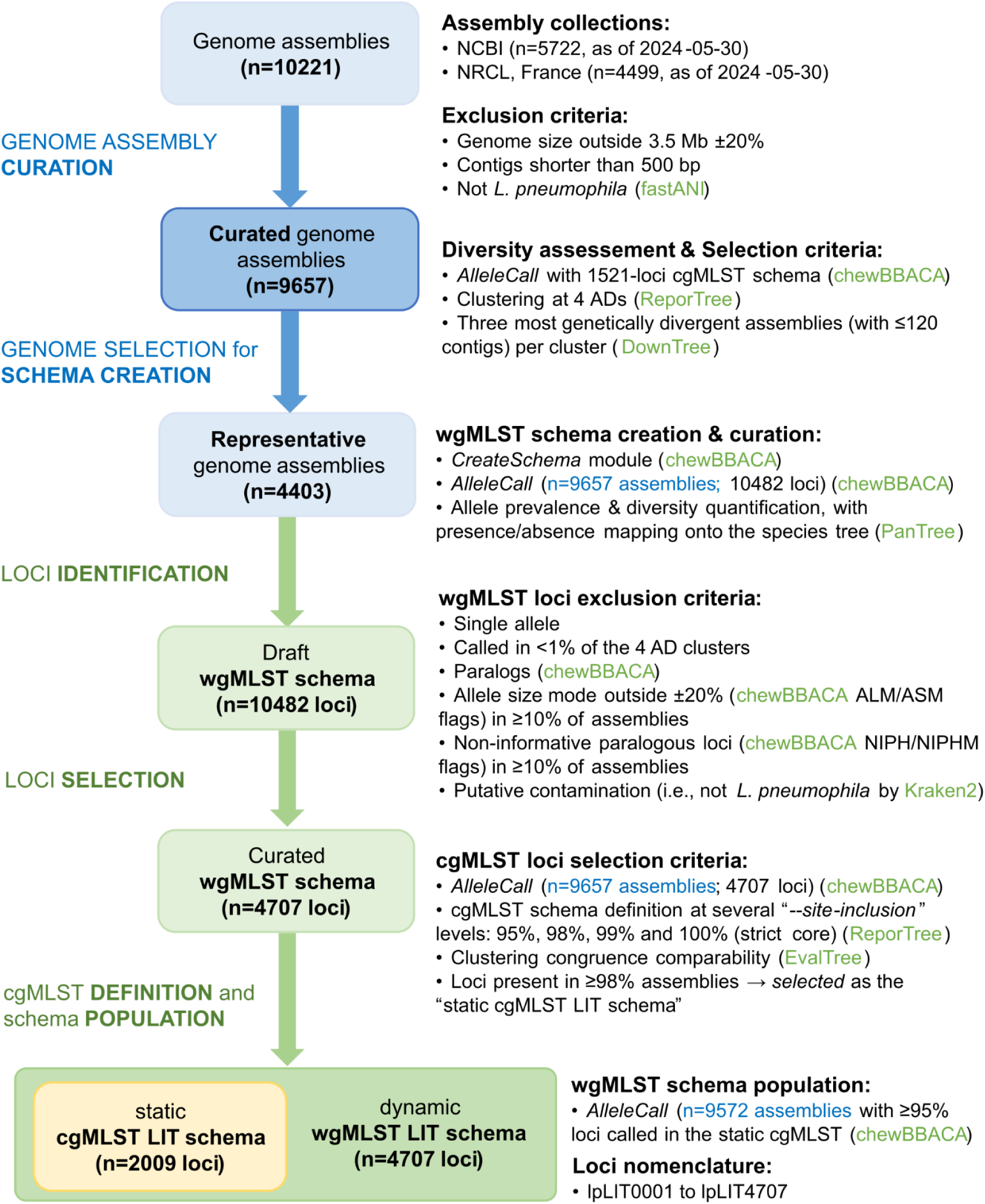
Schematic representation of the workflow used for the LIT cg/wgMLST schema creation, which is composed of 4707 loci, of which 2009 constitute the static cgMLST schema (Supplementary file S2).

**Figure 2.**
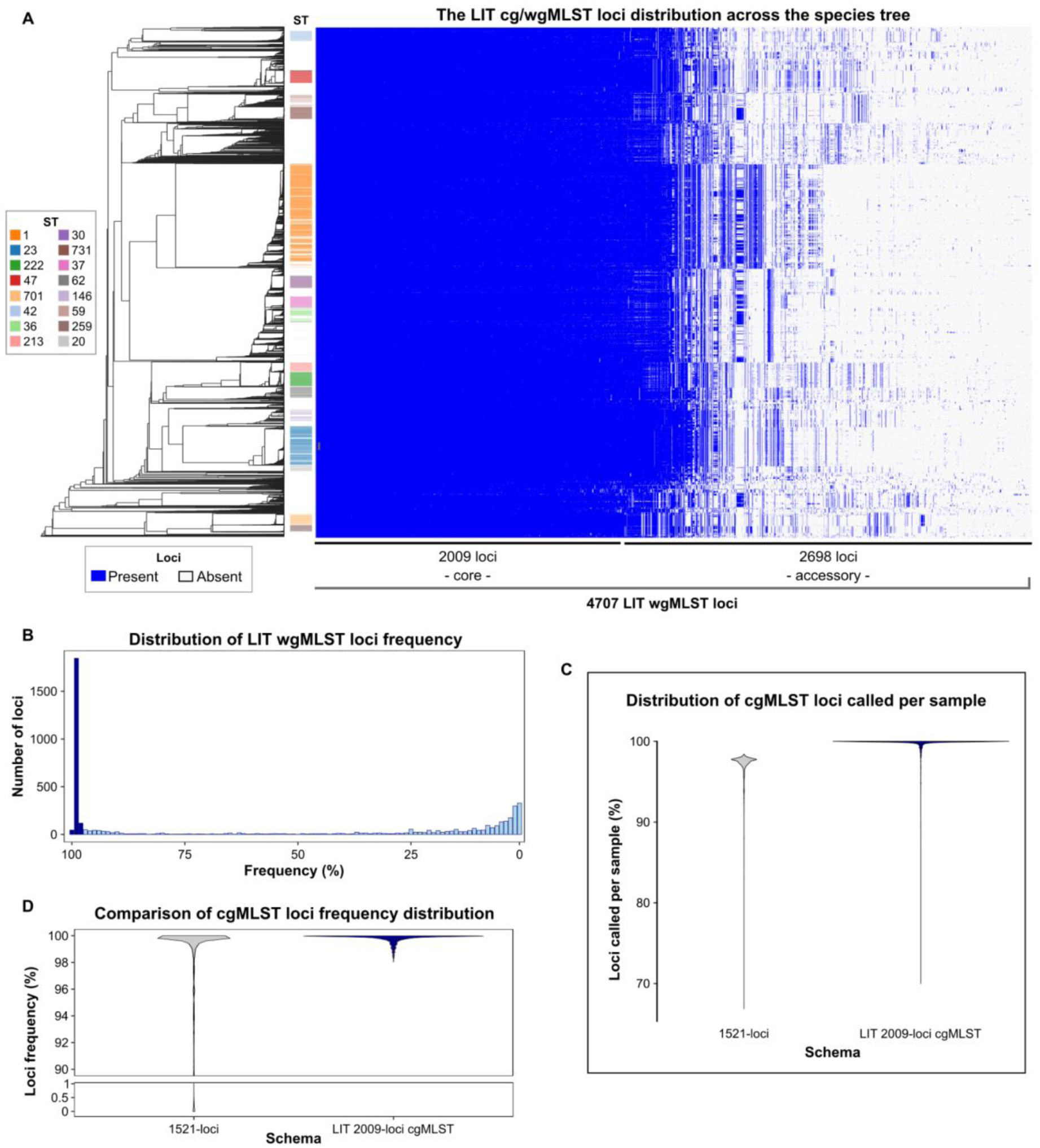
*Legionella pneumophila* LIT cg/wgMLST schema and comparison with the previously developed 1521-loci cgMLST schema. A) LIT cg/wgMLST loci distribution across the species tree (dendrogram generated with the 1521-loci schema), with indication of whether a locus is present (blue) or absent (white) in a given sample. Loci are ordered according to their prevalence in the dataset. Color blocks in the tips of the tree indicate the most prevalent STs in the dataset (≥100 samples each). B) Histogram indicating the number of loci present at each frequency in the dataset. The first bar (leftmost) corresponds to loci present in all samples (strict core), while subsequent bars show loci present in fewer samples, binned in intervals of 1% (e.g., [99%,100%[, [98%,99%[, etc.). C) Violin plot with the distribution of the loci frequency of the 1521-loci (n=1521) and the LIT (n=2009) cgMLST schemas. D) Violin plot with the distribution of the loci called per sample for the 1521-loci and the LIT cgMLST schemas (n=9657 genomes).

With the aim of defining a static cgMLST schema for routine genomic surveillance, different loci prevalence thresholds (95%, 98%, 99%, and 100%) were evaluated (Table 1). For comparison, the previously developed 1521-loci cgMLST schema [8] was similarly analyzed using a local chewBBACA deployment. Except for the strict core (100%), which clearly decreased the resolution power, all the LIT core schemas demonstrated higher resolution than the 1521-loci cgMLST, as reflected by an increased number of genetic clusters at the threshold of highest resolution of 0 ADs [8] (Table 1). Furthermore, the 1521-loci schema frequently yielded lower proportions of loci called per sample (median=97.6%; min=66.9%; max=98.4%; Figure 2C), resulting in more sample exclusion than any LIT core schema evaluated (Table 1). Among the different LIT core schemas evaluated, the 98% cgMLST represented the best compromise between resolution power and sample inclusion (Table 1), with more than 99% of genomes having at least 95% of loci called (median=99.95%; min=70.0%; max=100.0%; Figure 2C). Moreover, this cgMLST schema also presents better typeability than 1521-loci schema (Figure 2D). For this reason, the 2009 loci present in at least 98% of genomes were designated as the static cgMLST for the LIT schema (Figure 1 and Supplementary file S2), for which we suggest a threshold of ≥95% loci called for sample inclusion in clustering analysis, as subsequently applied in this study.

**Table 1.**
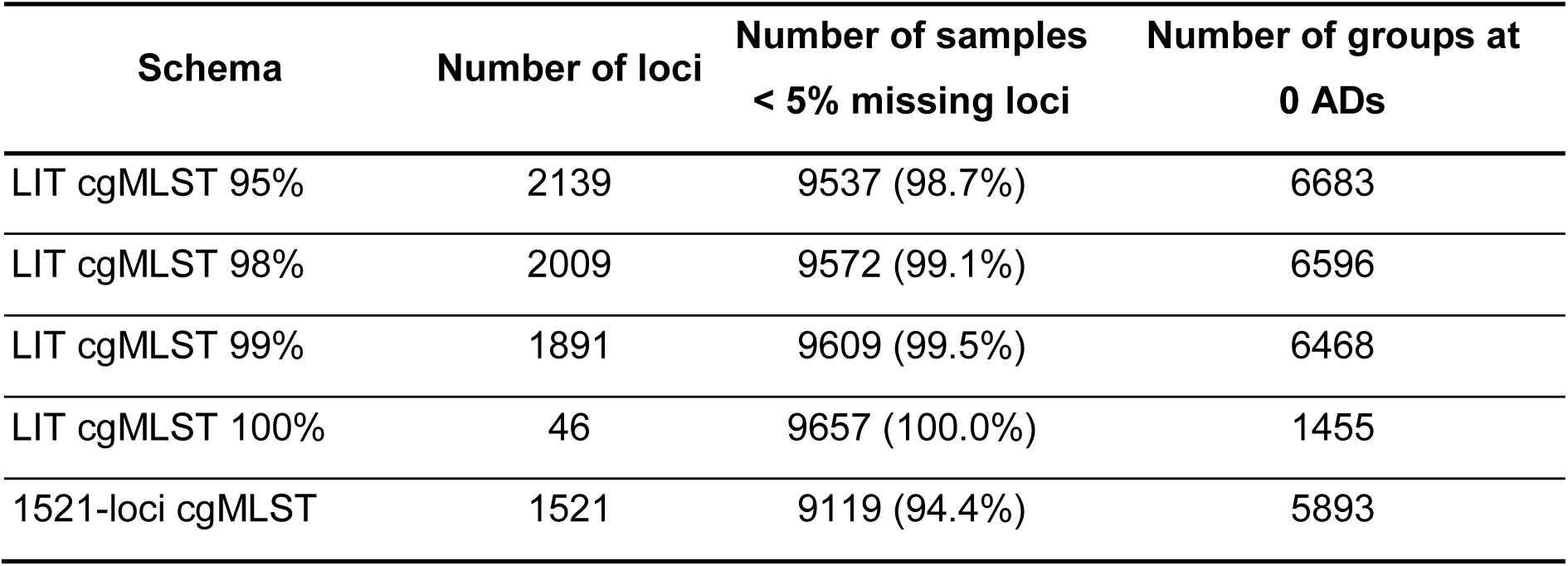
Evaluation of the different cgMLST schemas regarding the number of loci, the number and percentage of samples with less than 5% missing loci and the number of groups identified at 0 allelic differences (ADs), which was used as a proxy for resolution.

| **Schema** | **Number of loci** | **Number of samples**<br>**< 5% missing loci** | **Number of groups at**<br>**0 ADs** |
| --- | --- | --- | --- |
| LIT cgMLST 95% | 2139 | 9537 (98.7%) | 6683 |
| LIT cgMLST 98% | 2009 | 9572 (99.1%) | 6596 |
| LIT cgMLST 99% | 1891 | 9609 (99.5%) | 6468 |
| LIT cgMLST 100% | 46 | 9657 (100.0%) | 1455 |
| 1521-loci cgMLST | 1521 | 9119 (94.4%) | 5893 |

Ultimately, the LIT schema comprises a total of 4707 loci (2009 core and 2698 accessory) and has a total of 265193 unique alleles (median=48; min=1; max=694 per locus). This schema is available for download in Zenodo repository (https://doi.org/10.5281/zenodo.17871973) [33]. An adapted version of the schema is also available at chewie-NS (https://chewbbaca.online/) [34]. Of note, several loci of epidemiological/biological interest are included in the LIT schema, such as the genes *lag-1* (lpLIT0656) [35], *lpeA* (lpLIT2833), and *lpeB* (lpLIT2834) [36], which facilitates the rapid routine assessment of their presence/absence and intra-locus allelic diversity.

### Congruence with the 1521-loci cgMLST schema and backward compatibility with traditional typing

An important aspect of developing novel WGS-based typing methods is evaluating its clustering behavior relative to previously established typing approaches. From the 4707 loci that comprise the LIT wgMLST schema, 1436 loci (1337 core and 99 accessory loci) are among the loci that compose the 1521-loci schema [8] (Supplementary file S2). Using a recently applied cluster congruence assessment approach [24], an overall high congruence was observed between the two cgMLST schemas (Figure 3A and 3B and Supplementary file S6), indicating that the LIT static cgMLST captures a species population structure similar to that produced by a previous schema. Still, the LIT cgMLST schema yields a higher number of clusters when comparing identical AD thresholds, further showcasing its higher discriminatory power across all resolution levels (Table 1 and Figure 3B). For example, clusters detected by the 1521-loci cgMLST schema at 4 ADs, a threshold commonly used in this schema for signaling potential outbreak-related clusters [8,20], have the best correspondence to those detected at 6 ADs in the LIT cgMLST schema (CS=2.86; Supplementary file S6), reflecting the ability of the LIT schema to better resolve genetic diversity between closely related isolates. The performance of the LIT cgMLST schema was also benchmarked with another commonly used clustering algorithm (GrapeTree) [31]. The observed strong overall cluster congruence with single-linkage HC (Supplementary file S6) showcases the schema flexibility for implementation in most genomic surveillance workflows.

**Figure 3.**
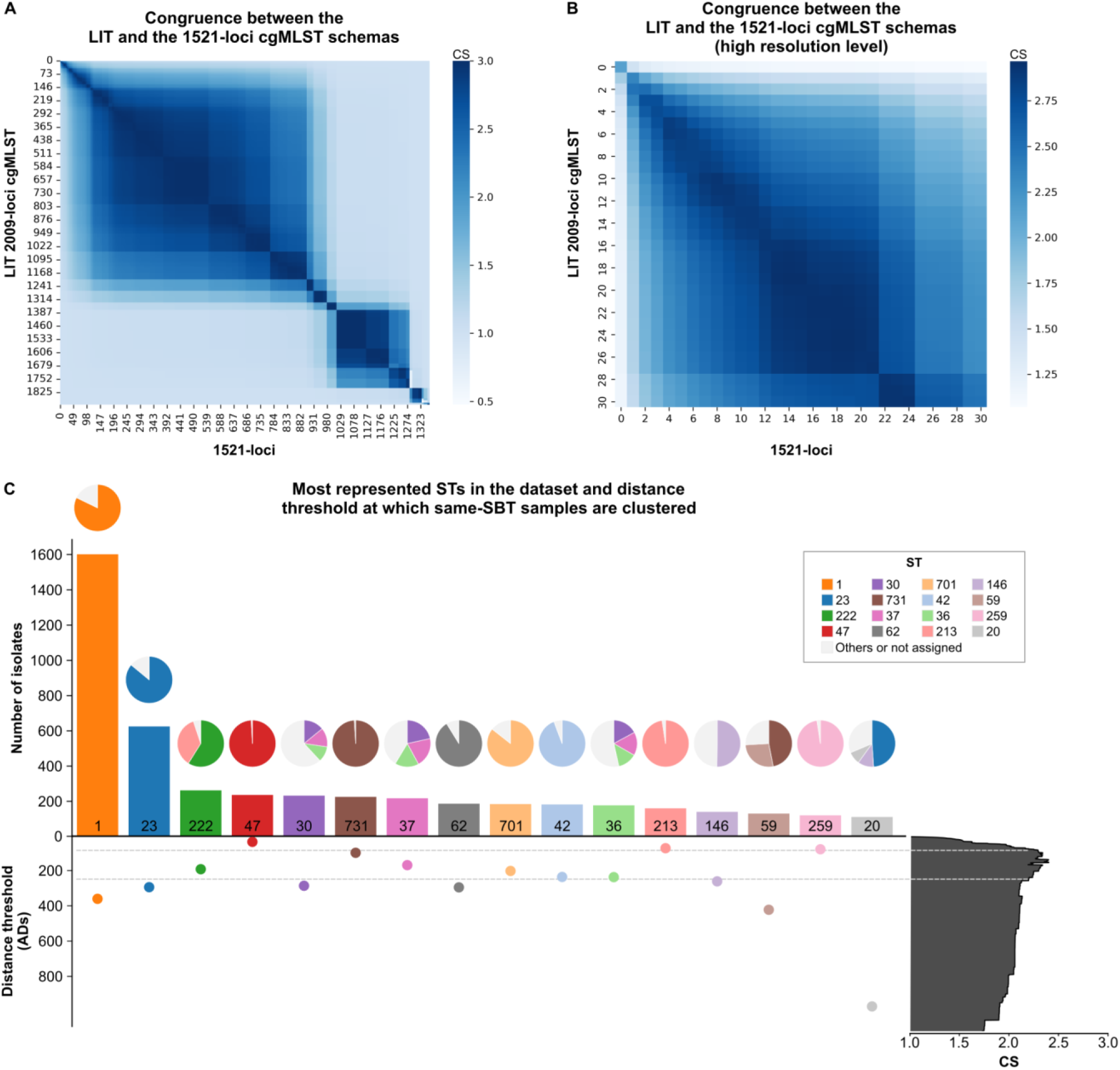
LIT cgMLST cluster congruence with the 1521-loci schema and backward compatibility with SBT. A) Heatmap of the cluster congruence score (CS) obtained from the comparison between the LIT cgMLST and the 1521-loci schema, at all possible resolution levels. B) Closer view of the heatmap presented in panel A at a high resolution level. C) Barplot with the number of samples per ST for the most represented STs in the dataset (≥100 samples each), accompanied by a swarmplot indicating the allelic distance (AD) threshold at which all samples of the respective ST are clustered together in the analysis with the LIT cgMLST schema. A piechart indicating the ST distribution in the cluster grouping all samples of a given ST is presented above the respective bar. The distribution of congruence scores (CS) derived from the evalTree analysis is shown on the right with CS threshold used to define high clustering congruence marked with a dashed line.

Regarding backward compatibility with *L. pneumophila* SBT classification (Supplementary file S1), the congruence assessment revealed that the single-linkage hierarchical clustering obtained at a threshold of 146 ADs with the LIT cgMLST schema has the best congruence with ST (CS=2.4; Figure 3C and Supplementary file S6). The fact that this CS is not close to the maximum possible (absolute congruence at CS=3) reflects not only differing levels of intra-ST genetic heterogeneity, but also the presence of STs that cannot be separated in a single cgMLST cluster (Figure 3C). Indeed, a closer inspection of the most represented STs in the dataset (≥ 100 samples each) revealed that the minimum AD threshold required to cluster all samples of an ST varies substantially across STs and often results in the merging of distinct STs (Figure 3C and Supplementary file S7). For instance, some STs (e.g. ST47 and ST731) had all samples clustered at AD thresholds considerably below the highest CS, while others (e.g. ST1, ST59 and ST20) revealed high intra-ST heterogeneity (Figure 3C). Besides informing about the backward compatibility with SBT classification during future LIT cgMLST adoption, these results clearly expose the high heterogeneity in genomic diversity of *L. pneumophila* STs at core genome level.

### The advantage of a dynamic wgMLST approach for *L. pneumophila* outbreak investigation

Clustering using core-genome loci revealed the presence of large clusters at high resolution levels, indicating that cgMLST alone might not be enough for outbreak investigation and source attribution. For instance, twelve of the clusters detected at 6 AD comprised more than 100 samples each (*i.e.*, together covering 22.8% the global dataset) (Figure 4A and Supplementary file S1). Given the non-uniform distribution of accessory loci across the *L. pneumophila* species tree, where distinct blocks of loci occur exclusively within specific branches (Figure 2A), the value of a dynamic wgMLST approach [22–24] was explored. With this approach, the static LIT cgMLST schema (n=2009 loci) is expanded on a cluster-by-cluster basis with accessory loci shared by all isolates within each cluster. Overall, when applied to all clusters identified at the 6 AD threshold (n=947), the dynamic wgMLST analyses incorporated a median of 616 additional accessory loci *per* cluster (Figure 4B and Supplementary file S8), thereby increasing the number of loci used for pairwise comparison within each cluster by an average of 30%. Specifically, the average number of loci used per cluster (median=2625; min=2230; max=2860) was close to the median number of wgMLST loci called per sample (median=2648; min=2343; max=2879), demonstrating that the dynamic wgMLST covers nearly the maximum number of loci that are comparable across same-cluster samples. Moreover, in total, 99.4% of the accessory loci of the LIT schema were used in the “zoom-in” analysis of at least one cluster (Supplementary file S8), further demonstrating the applicability of the whole schema. Notably, the dynamic expansion translated into increased confidence in the initial cgMLST-defined clustering, including, in many cases, a gain in discriminatory power (Figure 4C and 4D). Specifically, the same number of profiles was observed in 67.4% (638/947) of the cgMLST-defined clusters and an increased number of non-redundant allelic profiles was observed in 32.6% (309/947), resulting in improved confidence and isolate discrimination. In addition, the maximum intra-cluster pairwise allelic distance increased in 76.1% (721/947) of the clusters, further supporting that the inclusion of cluster-specific accessory loci enhances resolution and better captures the genetic diversity among closely related isolates.

**Figure 4.**
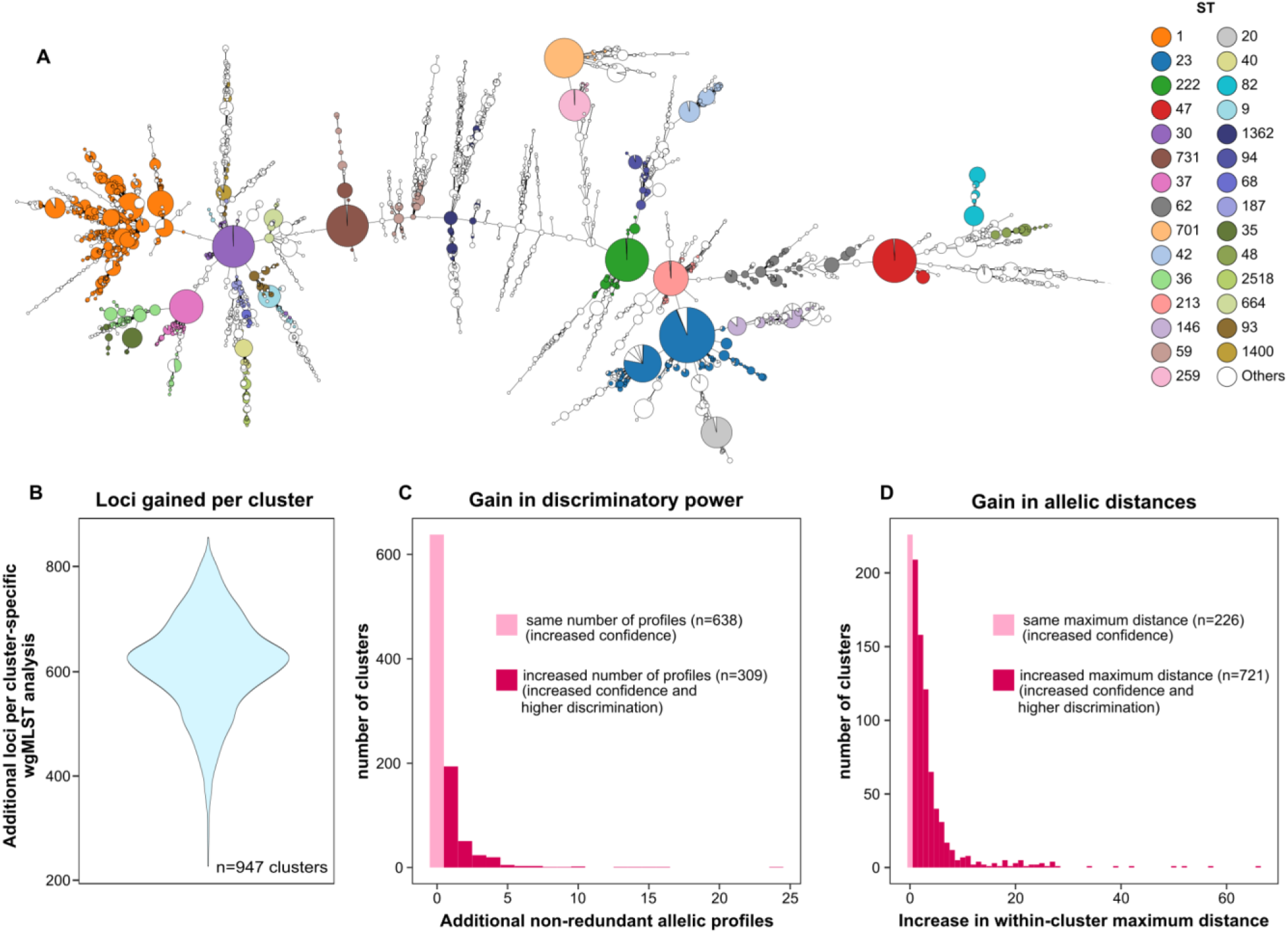
Cluster-specific dynamic wgMLST analysis using the LIT schema. A) Minimum Spanning Tree of the *L. pneumophila* dataset used in this study (obtained with the LIT cgMLST schema) with nodes colored by ST and collapsed at 6 allele differences, thus representing the cgMLST clusters used for zoom-in analysis. Branches are in log scale. B) Violin plot with the distribution of the number of accessory loci gained in cluster-specific dynamic wgMLST analysis (n=947 clusters). C) Histogram with the distribution of additional non-redundant allelic profiles gained per cluster. D) Histogram with the distribution of the maximum distance observed within each cluster. In panels B and C, light pink represents clusters in which no change was observed with the dynamic wgMLST analysis (increase confidence in cgMLST-based clustering), while dark pink represents clusters in which a higher intra-cluster discrimination could be achieved.

Given the high core-genome heterogeneity (Figure 3C) and differential accessory genome composition observed across STs (Figure 2A), all 6 AD clusters belonging to the most represented STs (n=330 clusters) were further examined to investigate whether an alternative strategy based on applying ST-specific cgMLST schemas, would be more suitable for such a diverse species. The data showed that, for each ST, the number of loci used in the dynamic zoom-in across all clusters, corresponding to the ST-specific core (*i.e*., the intersection of loci shared by every cluster of that ST), was consistently smaller than the total number of loci used when considering any cluster of the same ST (*i.e*., the union of loci present in at least one cluster) (Supplementary file S8). This pattern was particularly evident for the highly prevalent ST1, in which 1311 accessory loci were used in at least one cluster, whereas only 132 accessory loci were present in the ST-specific core. Together, these results indicate that, for some (if not all) *L. pneumophila* STs, designing multiple ST-specific cgMLST schemas would most likely yield lower resolution than the dynamic wgMLST approach proposed in this study.

To evaluate the practical implications of this dynamic wgMLST approach in a real-life surveillance context, nine groups of epidemiologically-related *L. pneumophila* isolates (referred as “investigations”) corresponding to eight different STs were analyzed with the entire dataset (Figure 5 and Supplementary file S8). The performance of the dynamic wgMLST LIT schema was compared with that of the static cgMLST LIT schema using metrics assessing cluster delineation, separation, discrimination and confidence (Figure 5). As expected, the static cgMLST effectively served as a first-line detection of clustering signal, correctly grouping same-investigation isolates within the same 6 AD cgMLST cluster for all investigations (9/9). Of note, none of these clusters were exclusive to the investigation isolates, suggesting insufficient resolution of the static cgMLST to clearly delineate outbreak boundaries, although it was still able to subcluster the isolates in four investigations (Supplementary file S8). Besides the inherent gain in confidence obtained by relying on a substantially higher number of loci, the subsequent application of a dynamic wgMLST strategy to each cluster also resulted in gains in at least one of the other three metrics for seven out of nine investigations (Figure 5 and Supplementary file S8). For instance, the dynamic wgMLST successfully increased the allelic distance between investigation and non-investigation isolates in six out of nine investigations and the intra-investigation discrimination (*i.e.*, higher number of non-redundant allelic profiles) in three, demonstrating that this approach reveals subtle genetic variation that would remain undetected by static cgMLST alone. Notably, in all but one investigation (investigation 5) where no apparent gain in intra-investigation discrimination was observed, the maximum SNP distance between the investigation isolates and the reference (one of the clinical isolates) was comparable to the maximum AD obtained by wgMLST, differing by no more than one unit (Figure 5 and Supplementary file S8). This indicates that maximum, or near-maximum, resolution was achieved using cluster-specific dynamic wgMLST. Overall, this real-life practical exercise illustrates the flexibility, consistency and robustness of the dynamic wgMLST approach for isolates’ discrimination and source attribution in *L. pneumophila* outbreak and environmental investigations.

**Figure 5.**
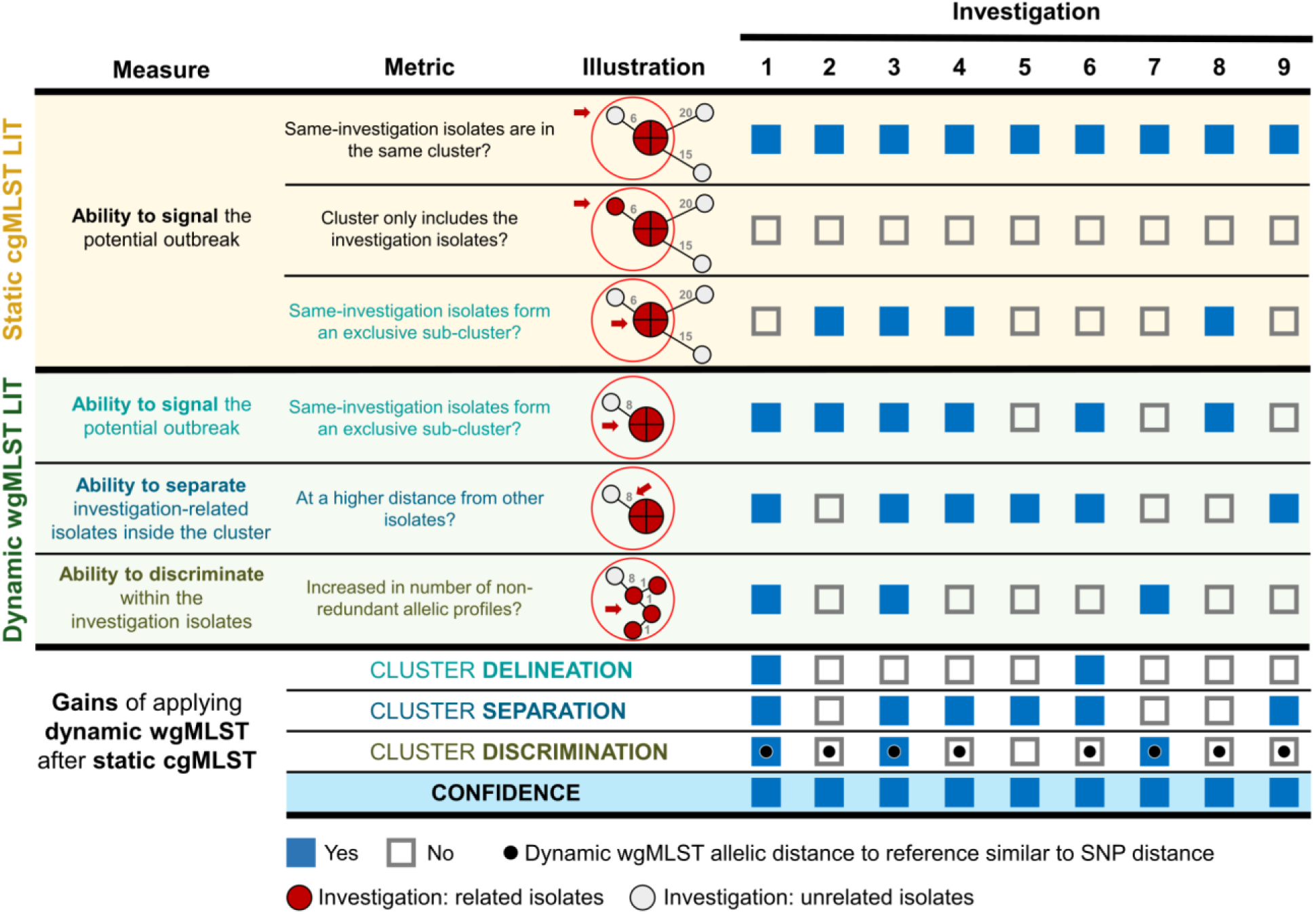
Assessment of the resolution gains of applying a dynamic wgMLST after a static cgMLST analysis for nine groups of epidemiologically-related *L. pneumophila* isolates (referred as “investigations”). Three main metrics (cluster delineation, cluster separation and cluster discrimination) were evaluated based on objective Yes/No questions for both the initial static cgMLST clustering (yellow background) and the subsequent dynamic wgMLST clustering (green background) applied to each cgMLST 6 AD cluster containing isolates from the same investigation. For each question, an illustration is provided to demonstrate what is being assessed in the clustering tree, as highlighted by red arrow and circle. For each investigation, the bottom overview panel presents an objective Yes/No assessment of whether the dynamic wgMLST translated in gains for each metric, as well as for the overall cluster confidence, defined as the strength of genetic evidence based on gains in at least one of the previous three metrics and/or in the number of loci supporting the investigation results. Black dots in the ‘cluster discrimination’ squares indicate investigations in which the dynamic wgMLST approach achieved resolution comparable to reference-based SNP analysis, defined as cases where the maximum allelic distance between investigation isolates and the reference differed by no more than one unit between the two approaches. Details on this analysis are provided in Supplementary file S8.

### Cluster stability regions and perspectives for future nomenclature design

The implementation of cgMLST-based hierarchical nomenclature systems is increasingly being adopted in genomic surveillance [37–39], as it facilitates continuous tracking of circulating genogroups and supports outbreak investigations. Cluster stability regions are defined as contiguous AD thresholds over which cluster composition remains consistent, thus constituting good targets for nomenclature design. Several stability regions were found within the LIT cgMLST schema across different levels of resolution (details in Supplementary file S5). None of these regions overlapped with the threshold presenting highest congruence with SBT classification (146 ADs), suggesting that this threshold is inadequate for a stable cgMLST-based nomenclature. Still, three large stability regions were found with mid threshold points around 1500, 600 and 400 ADs, being candidate *plateau* levels for the implementation of a novel nomenclature that reflects species population structure. Additionally, two stability regions at higher resolution levels were identified (∼80 and ∼25 AD thresholds; Supplementary file S5), unveiling the suitability of the LIT static cgMLST schema for the implementation of a comprehensive high-resolution hierarchical nomenclature for longitudinal surveillance of circulating genogroups.

## Discussion

The development and release of the LIT cg/wgMLST schema aimed to advance global *L. pneumophila* genomic epidemiology by offering improved resolution, representativeness, and flexibility compared with previously available typing methods. By relying on over 9600 genome assemblies, the LIT schema, comprising 4707 loci, of which 2009 constitute the static cgMLST component (defined by ≥98% prevalence across the global dataset), captures a much wider spectrum of *L. pneumophila* genomic diversity than earlier cgMLST schemas [8,9]. In local deployments, the LIT cgMLST demonstrates higher typability than the 1521-loci schema [8], as evidenced by a greater proportion of loci called per genome, enabling broader sample inclusion in clustering analyses (Figure 2D). It also provides consistently higher resolution, as demonstrated by the identification of a greater number of clusters when comparing similar AD thresholds (Table 1), including the threshold commonly employed for outbreak detection and investigation [8,20].

Besides these gains, the LIT schema addresses the challenges associated with genomic typing of a highly diverse bacterial species with an open pangenome, such as *L. pneumophila*, by providing a dual framework: i) a static cgMLST for standardized comparison in routine scenarios, and ii) a dynamic wgMLST for high-resolution “zoom-in” analyses of cgMLST clusters of interest, following a previously described rationale [22–24]. This approach reconciles reproducibility with flexibility, with the static cgMLST serving as a first-line screening tool to identify potentially linked isolates, followed by an optional dynamic and context-adaptive schema expansion with cluster-specific accessory loci to refine outbreak delineation. The observed gains in discriminatory power, and consequently in genetic evidence of isolate linkage, were clearly demonstrated through comprehensive large-scale dynamic wgMLST assessments at potential outbreak level. For example, the dynamic approach increased discriminatory capacity in over 30% of the 947 assessed clusters and revealed additional intra-cluster diversity in more than 70% of cases, uncovering fine-scale variation undetectable by the static cgMLST schema alone (Figure 4B and 4C). These gains were further evidenced in the analysis of epidemiologically-related isolates, where the dynamic wgMLST enhanced the confidence for outbreak detection and source attribution through gains in cluster delineation, separation and discrimination. Notably, wgMLST achieved maximum, or near-maximum, resolution in most of the investigations with wgMLST, capturing distances of the same magnitude of reference-based SNP analysis against a cluster representative sequence. In this context, the cluster-specific dynamic wgMLST approach implemented in the LIT schema emerges as an alternative to traditional SNP-based analysis, which requires the selection of a cluster-representative reference sequence and the parallel analysis with a different and computationally demanding bioinformatics workflow, involving read mapping, variant calling and advanced phylogenetics [40]. By contrast, the operationalisation of dynamic wgMLST is considerably less laborious and more straightforward, as the initial cluster detection and subsequent in-depth investigation rely on the same input matrix (*i.e*., the wgMLST allelic matrix). Overall, the scalability of cg/wgMLST makes it particularly well suited to both local and global surveillance frameworks, where harmonized typing and interoperable data structures are essential [41–43]. Supporting its seamless implementation within national reference laboratories and international genomic surveillance networks, the LIT schema is fully compatible with the widely used allele caller chewBBACA [26], already adopted by national and international entities such as the ECDC and the European Food Safety Authority (EFSA) [44]. Moreover, the two-tier strategy behind the dynamic wgMLST can be automated with the “zoom-in” functionality provided by ReporTree [23], as performed in this study. A comprehensive guide for schema implementation and operationalisation is provided together with the schema in the Zenodo repository (https://doi.org/10.5281/zenodo.17871973) [33].

A key consideration in transitioning from established methods such as SBT to WGS-based typing is the preservation of continuity with historical classifications. Our assessment of backward compatibility revealed a moderate concordance between cgMLST clusters and traditional STs (Figure 3C). This became even more evident when examining in greater detail the most represented STs in the dataset, which showed substantial heterogeneity across both core and accessory genomes (Figures 2A and 3C), with several prevalent STs, including ST1, displaying pronounced intra-ST diversity. Overall, this partial overlap between ST-based classification and cgMLST clustering was not unexpected [10,45–47]. This reflects the well-recognized limitations of the seven-locus SBT scheme for *L. pneumophila* [8–11], and possibly the underlying evolutionary dynamics of the species, in which homologous recombination plays an important role in shaping population structure and evolution [46,47]. In this context, while recombination poses challenges for any genomic analysis, particularly when inferring genetic relatedness, cgMLST offers important advantages for routine monitoring of circulating genomic diversity. Specifically, allele-based distance metrics can buffer the impact of recombination, preventing isolates that share a recent common ancestor from appearing artificially distant. Furthermore, our evaluation of the proposed dynamic wgMLST approach at the ST level indicates superior resolution and usability for *L. pneumophila* when compared to ST-specific cgMLST schemes. Indeed, the application of ST-specific schemas or a cumulative cgMLST approach (as performed for other species [48]) would be substantially more challenging in terms of design and operationalization, as it would require multiple schemas that are unlikely to comprehensively capture all epidemiologically relevant STs, while offering limited flexibility and being poorly suited to the high diversity and only moderate concordance observed between ST and cgMLST-based clustering. It is important to stress that the exercises for detecting potential outbreak signals using the LIT schema applied a single cluster threshold (6 ADs, as it showed better cluster agreement with the 4 AD threshold traditionally used with the 1521-schema), but outbreak case definition based on WGS data always requires tailoring genetic thresholds to the specific context of each investigation. Genetic and temporal distances among potentially epidemiologically linked isolates, the expected genetic heterogeneity and available typing information of the “strain” under investigation should be considered among other indicators [24]. Our data indicates that this is particularly necessary for *L. pneumophila*, where pronounced differences in genetic diversity at both intra- and inter-ST levels are evident, undermining the application of a single outbreak-level threshold across the species, which would likely inflate “outbreak” clusters for highly clonal STs. Further work is needed to inform threshold ranges that maximize epidemiological concordance.

The adoption of standardized genome-based cluster nomenclature systems, such as hierarchical nomenclatures or Life Identification Numbers [43,48–50], represents a logical next step to ensure global interoperability and consistent communication of *L. pneumophila* genomic relationships obtained with the LIT schema. The identification of multiple clustering stability regions, from broad population structure thresholds (∼1500, 600 and 400 ADs) to genogroup-level resolution (∼80 and ∼20 ADs) provides a first and robust empirical basis for future nomenclature design with the LIT schema, supporting both high-resolution outbreak investigation and long-term lineage tracking within routine surveillance programs. While the implementation of a globally agreed nomenclature framework requires further inter-laboratory and international collaborative work, at the local level, the establishment of multi-level hierarchical nomenclatures can already be straightforwardly operationalized, again taking advantage of the availability of such a feature in tools like ReporTree [23]. Noteworthy, despite being fully tailored for implementation with chewBBACA allele caller [26] followed by cg/wgMLST hierarchical clustering analysis and local nomenclature with ReporTree [23], the LIT schema can be implemented in workflows using different software and algorithms, provided that a prior evaluation of loci typeability and clustering congruence is conducted as for any other schema [24], as exemplified in this study by the HC- and GrapeTree-based comparison (Supplementary files S5 and S6). These implementation considerations should be taken into account in future integration of the LIT schema into public databases and open-source analysis platforms (e.g. PubMLST, https://pubmlst.org/ [51]), which have the potential to enhance inter-laboratory comparability, as well as facilitate continued schema expansion and the implementation of an internationally established cluster nomenclature as new genomic diversity is uncovered. Still, sustained contributions will remain essential to ensure long-term stability, transparency, and global adoption of this typing framework. Indeed, the previous experience and success of the global adoption of SBT demonstrated how critical these aspects are, while also highlighting the need for a more cooperative, globally coordinated curation model, ideally supported by stakeholders that can ensure long-term sustainability. As WGS is increasingly implemented in surveillance, the improved and standardized genomic approach provided by the LIT schema could help reduce disparities in data analysis and reporting, ultimately leading to more effective responses and lower overall costs [1,52]. In addition to its value for *L. pneumophila* genomic surveillance, the wgMLST resources generated are likely to accelerate research and bridge routine surveillance with scientific investigation, by offering a versatile framework to screen the presence/absence and allelic diversity of genes with biological relevance (e.g. the *lag-1*, *lpeA*, and *lpeB* intentionally included in the schema). This enables practical applications, such as rapid genome characterization and the development of targeted screening of genetic determinants underlying ecological success and pathogenicity.

In summary, the LIT cg/wgMLST schema represents a comprehensive, scalable, and backward-compatible framework that advances the genomic surveillance of *L. pneumophila*. When coupled with epidemiological metadata, the LIT-based cg/wgMLST can enable real-time detection of emerging lineages, cross-border cluster tracing, and linkage of clinical and environmental isolates, capabilities essential for managing *Legionella* outbreaks, which often involve complex exposure histories and delayed source identification [53,54]. By combining a robust static cgMLST schema with a flexible, dynamic wgMLST schema, this cg/wgMLST approach balances standardization and high-resolution adaptability, providing superior capacity to resolve closely related isolates and overcoming key limitations of both SBT and previously developed cgMLST schemas [8,9,14]. Finally, its enhanced discriminatory power, and readiness for integration into hierarchical nomenclature systems position it as a key tool for *Legionella* surveillance. Through continued refinement and global coordination, this framework can substantially contribute to harmonized, genome-informed monitoring of Legionnaires’ disease at both national and international scales.

## Statements

### Ethical statement

Ethical approval was not necessary because the data used for the study were collected as part of the infectious disease surveillance defined by national legislations. All data was processed anonymously.

### Use of artificial intelligence tools

None declared.

### Data availability

Accession numbers for NCBI genome assemblies are detailed in Supplementary file S1. The LIT cg/wgMLST schema, and associated resources for immediate applicability for standardized, high-resolution WGS-based routine genomic surveillance, are publicly available at Zenodo repository (https://doi.org/10.5281/zenodo.17871973) [33]. An adapted version is also available at chewie-NS (https://chewbbaca.online/). Supplementary material generated in this study is also available in Zenodo (https://doi.org/10.5281/zenodo.18473692). Investigation-associated isolates raw reads are available on ENA under BioProject accession number PRJEB104302.

## Acknowledgements

The authors are grateful to Baharak Afshar and Rediat Tewolde (UKHSA), for their support and assistance in establishing the *Legionella* International Typing (LIT) workgroup. We would also like to thank the LIT workgroup for their support and intellectual suggestions including Melina Bigler, Teresa Fasciana, Valeria Gaia, Anna Giammanco, Tim Handschin, Marit Hetland, Laurine Kieper, Anne Krøvel, Daniel Mäusezahl, Maria Scaturro, and Jennifer Tanner. We also thank ECDC colleagues including Lara Payne Hallstrom, Marina Papamichael, Maximilian Riess, and Olov Svartstrom. Lastly, the authors thank Mário Ramirez and Rafael Mamede from Faculty of Medicine of the University of Lisbon (FMUL) and Gulbenkian Institute for Molecular Medicine (GIMM) for their kind support in releasing the LIT wgMLST schema in Chewie-NS (https://chewbbaca.online/).

